# Preserved C-reactive protein responses to blood stream infections following tocilizumab treatment for COVID-19

**DOI:** 10.1101/2021.07.03.21259949

**Authors:** Emmanuel Q Wey, Clare Bristow, Aarti Nandani, Bryan O’Farrell, Jay Pang, Marisa Lanzman, Shuang Yang, Soo Ho, Damien Mack, Michael Spiro, Indran Balakrishnan, Sanjay Bhagani, Gabriele Pollara

## Abstract

C-reactive protein (CRP) levels are elevated following bacterial infections but may be attenuated by the IL-6-receptor antagonist tocilizumab. In hospitalised COVID-19 patients, tocilizumab induced a transient (<21 day) fall in CRP but retained CRP responses to nosocomial blood stream infections, and therefore its utility in guiding antibiotic prescribing.

## Introduction

Bacterial co-infection in COVID-19 is infrequent and challenging to exclude [1,2], but occurs more commonly in longer hospital stays and following admission to the intensive care unit (ICU) [3]. This dichotomy emphasises the role of biomarkers to both diagnose bacterial co-infection, guiding appropriate antibiotic use, as well as support withholding unnecessary antibiotics to minimise selection for antimicrobial resistance.

Elevations in C-reactive protein (CRP) concentration are commonly taken to indicate bacterial infections, but many factors promote secretion of this acute phase response protein, including increases in circulating IL-6 cytokine [1]. Severe COVID-19 is characterised by a hyperinflammatory state partly driven by elevated IL-6 activity, as demonstrated by the mortality benefit offered by tocilizumab, a humanised monoclonal antibody targeting the IL-6 receptor [4,5].

One consequence of tocilizumab-mediated inhibition of the IL-6 signalling pathway is that routinely measured CRP concentrations are decreased [6]. In inflammatory arthritides, tocilizumab doses are administered every 2-4 weeks, resulting in variable attenuation of CRP responses to bacterial infection [7–9]. In COVID-19, single doses of tocilizumab are used [4,5], making the extent and duration of IL-6 signalling suppression less defined [10]. In a small COVID-19 cohort with blood stream infections (BSIs) that had received tocilizumab, CRP concentrations were reduced at the time of BSI diagnosis, but remained above normal values [11]. However, kinetics of the CRP response around the onset of BSI were not assessed, and thus it remains unknown whether tocilizumab use in COVID-19 attenuates the utility of CRP to guide antibiotic prescribing [1,12]. We sought to address this research question by identifying a cohort of COVID-19 patients receiving a single dose of tocilizumab and testing the hypothesis that tocilizumab use in this context did not impair elevations in CRP in response to bacterial infections, as modelled by BSIs.

## Methods

### Data extraction and ethics

Anonymised demographics, tocilizumab and corticosteroid prescriptions, ICU admissions, haematological and biochemical investigations were extracted as previously described [1]. The study was approved by the Research and Innovation Group at RFL NHS Trust, which stated that as this was a retrospective review of routine clinical data, formal ethics approval was not required.

### Patient selection

All patients included in this study were admitted at the Royal Free Hospital (RFH), London between 1 March 2020 and 1 February 2021 inclusive. We included patients aged >18 years admitted to hospital with a COVID-19 diagnosis defined by either RT-PCR detection of SARS-CoV-2 from nasopharyngeal swabs or a compatible clinical and radiological syndrome. Patient use of tocilizumab originated either through delivery of routine clinical care or from randomised clinical trials after unblinding. COVID-19 associated blood stream infections (BSI) were defined as isolation in blood cultures of any bacteria, excluding coagulase negative staphylococci, between 14 days prior to and 60 days after a COVID-19 diagnosis was made. We excluded patients that developed BSIs prior to receipt of tocilizumab. To assess dynamic CRP responses, we included only patients with routine blood measurements at least 3 days prior to onset of BSI.

### Statistical analysis

Baseline demographics were compared by Mann–Whitney test (age), Fisher’s exact test (gender and microbiology) or Chi-square test (ethnicity and Charlson co-morbidities). Continuous variables were expressed as median and IQR, and patient groups were compared using non-parametric two-tailed Mann–Whitney tests. All analyses were performed using Microsoft Excel and GraphPad Prism.

## Results

We identified 107 patients admitted to RFH with COVID-19 who received tocilizumab as part of routine clinical care or within the context of randomised clinical trials. Of these individuals, 17 developed a BSI during their hospital admission (table S1). We also identified a comparator group of 55 patients with COVID-19 who did not receive tocilizumab but that developed a BSI during their hospital admission (table S1). Tocilizumab use was more commonly associated with ICU admission, but the range of organisms responsible for BSI was comparable between the groups (table S1).

To assess the impact of tocilizumab on inflammatory marker dynamics, we derived CRP concentrations and white cell counts over time in tocilizumab recipients. As expected, CRP fell rapidly in the first week after tocilizumab administration (fig 1A), whereas this was not observed for total white cell, neutrophil or lymphocyte counts (fig 1A & fig S1). The reduction in CRP following tocilizumab was short lived, however, patients showing a rise in CRP concentration within 21 days of receiving the drug (fig 1A). To exclude confounding by bacterial co-infection, we performed a sensitivity analysis on 90 patients that did not develop a BSI following receipt of tocilizumab, and an early reduction followed by rebound in CRP was still observed (fig S2A). A similar pattern was evident in patients that developed a BSI, although notably CRP concentrations showed less attenuation and greater heterogeneity within the 21 day period since tocilizumab administration (fig S2B).

**Figure 1.**
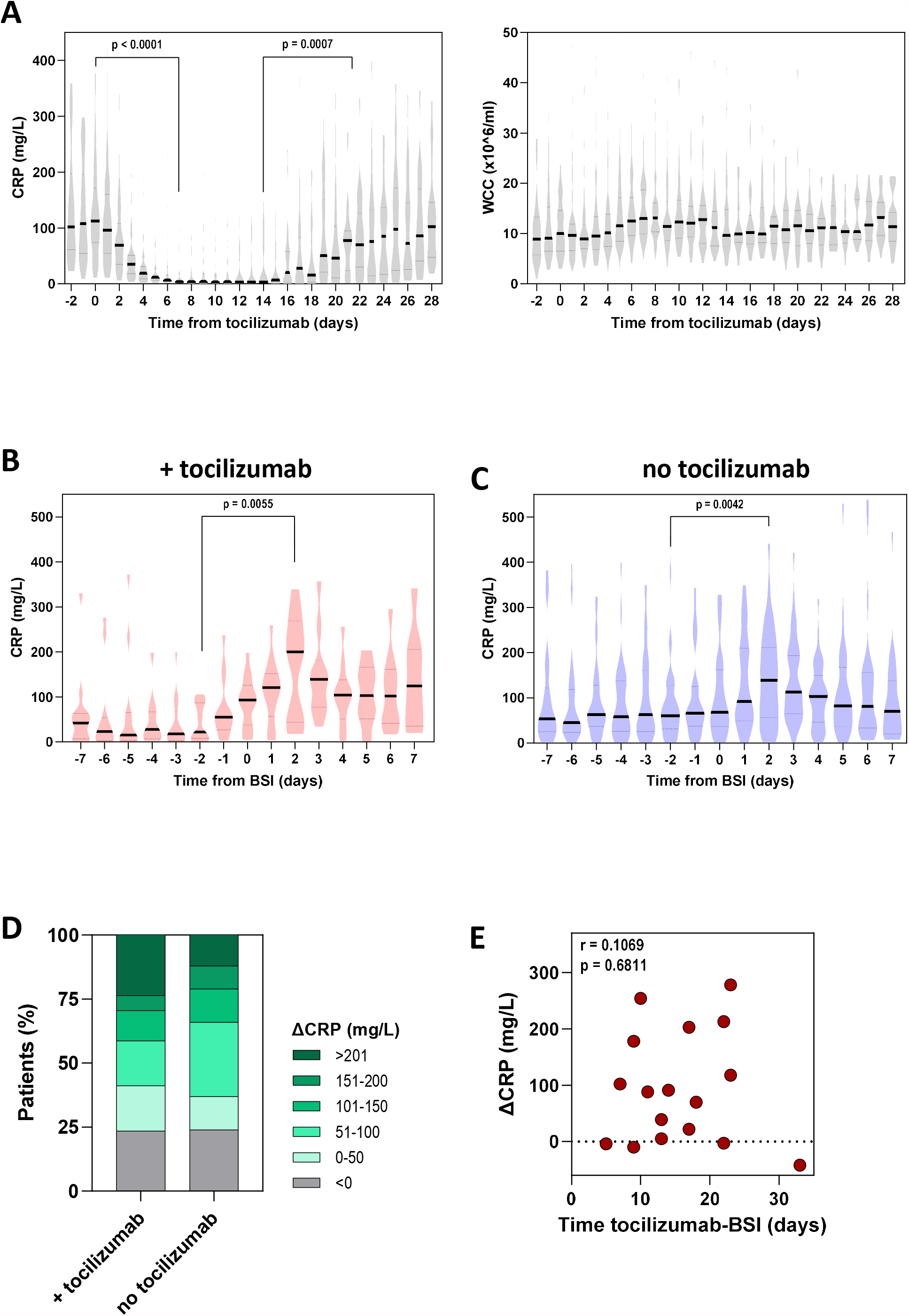
Inflammatory marker responses following tocilizumab administration and onset of BSI in COVID-19. (A) CRP concentration (left panel) and white cell count (right panel) relative to time since tocilizumab administration. All COVID-19 patients receiving tocilizumab included in analysis, independent of a subsequent presence of BSI (n = 107). (B) CRP concentration over time in patients that received tocilizumab and subsequently developed a BSI (n = 17). (C) CRP concentration of time in COVID-19 patients that developed a BSI but with no prior tocilizumab exposure (n = 55). (D) Change (Δ) in CRP induced by onset of BSI in COVID-19 patients stratified by prior receipt of tocilizumab. (E) Relationship between ΔCRP induced by BSI and time between administration of tocilizumab and the onset of BSI. Violin plots represent frequency distribution of all samples, with bold and dashed lines representing median and quartile values. All p values were calculated by Mann-Whitney tests.

Next, we investigated whether the tocilizumab-mediated reduction in CRP negated the diagnostic utility of this biomarker during an episode of BSI in COVID-19. To test the hypothesis that CRP concentrations would rise following a BSI independent of prior tocilizumab administration, we compared CRP responses in 17 patients who had received tocilizumab prior to a BSI with 55 patients who had not received tocilizumab. Strikingly, in both cohorts, the onset of a BSI resulted in a clear elevation in CRP (figs 2B & 2C). To determine whether the CRP elevation was quantitatively comparable, we calculated the change in CRP across the time of BSI onset. As blood samples were not universally collected daily, we derived paired sampling for all patients by calculating the maximal CRP value 2 or 3 days prior to blood culture sample collection and comparing this to the maximal CRP up to 2 days after BSI. This approach revealed an increase in CRP following BSI in 76.5% and 75.0% of patients that had or had not received tocilizumab respectively (fig 2D). Moreover, there was no quantitative difference in CRP increase between the groups (median CRP change +88 mg/L vs +76 mg/L respectively, p = 0.67 by Mann-Whitney test).

Finally, as patients developed BSIs at varying times following receipt of tocilizumab, we tested the hypothesis that BSI-induced CRP increment would be proportional to the time interval between tocilizumab administration and BSI onset. However, in the 17 patients that both received tocilizumab and subsequently developed a BSI, no relationship was observed between the size of the tocilizumab-BSI interval and the change in CRP (r = 0.1069, p=0.6811 by Rank-spearman correlation) (fig 2E).

## Discussion

Tocilizumab attenuates IL-6 signalling, removing one of the drivers for CRP secretion, a pivotal tool in the diagnosis and exclusion of bacterial infections, including in COVID-19 [1,12]. Tocilizumab use to improve patient outcomes in severe COVID-19 could have the unintended consequence of removing the ability of CRP to guide judicious antibiotic prescribing [4,5]. However, we demonstrate that prior administration of a single dose of tocilizumab does not attenuate CRP responses following the onset of a BSI, retaining the clinical utility of this diagnostic biomarker in this context.

Preservation of CRP responses following tocilizumab has important implications for the care of patients with severe COVID-19. First, false reassurance from the lack of CRP increments in confirmed BSIs occurs no more frequently than in the absence of tocilizumab, reducing the risk that tocilizumab use would delay the prescription of indicated antibiotics. Second, low CRP levels in tocilizumab recipients is not an indication for continued prescription of unnecessary antibiotics, supporting antimicrobial stewardship efforts in this cohort of patients. Nevertheless, it was notable that BSI onset did not initiate CRP elevations in all patients, independent of prior tocilizumab use, emphasising that CRP should be only one of several variables that are used to diagnose incipient bacterial infections.

Despite preserved CRP responses to BSI, tocilizumab use did reduce baseline CRP concentrations. However, this effect was transient, CRP levels recovering at varying times, mostly within 3 weeks. In addition, increments in CRP after BSI onset were not related to the time since tocilizumab administration, indicating that CRP increment is unlikely to be solely IL-6 dependent or that in COVID-19 neutralisation of the IL-6 axis by a single tocilizumab dose is not absolute. Measuring IL-6 signalling activity in vivo may predict attenuation of CRP responses in some individuals, as well as inform the need for further tocilizumab dosing in COVID-19 [10].

Our study has limitations. The single-centre, retrospective source of our clinical data constrained numbers of patients that could be identified in each group, negating the ability to correct for potential confounders. Nevertheless, the increased frequency of corticosteroid use in patients with a BSI that had received tocilizumab may have attenuated CRP responses further, counter to our observations. BSIs provided a standardised, prototypic model for bacterial infections to compare across clinical groups, but limited extrapolation to non-BSI settings. Further studies of COVID-19 associated non-BSI bacterial co-infections will be needed to confirm the generalisability of our findings.

In conclusion, we show that tocilizumab use in severe COVID-19 does not prevent elevations in CRP concentration following the onset of a confirmed bacterial co-infection, as modelled by BSIs. Use of tocilizumab should not negate judicious, CRP-guided use of antibiotics in COVID-19.

## Data Availability

All data analysed in the manuscript are depicted in the figures. Anonymised raw data are available upon request from the corresponding author.

---

**Table S1.** Baseline demographics and clinical characteristics for patients included in the study.

|  | No BSI & received<br>tocilizumab<br>(n = 90) | BSI & received<br>tocilizumab<br>(n = 17) | BSI & did not receive<br>tocilizumab<br>(n = 55) | p value* |
| --- | --- | --- | --- | --- |
| **Age**, years, median (range) | 63 (28-90) | 61 (50-78) | 63 (31-100) | p = 0.383 |
| **Gender**, n (%) |  |  |  |  |
| Male | 56 (62) | 9 (53) | 38 (69) | p = 0.253 |
| Female | 34 (38) | 8 (47) | 17 (31) |  |
| **Ethnicity**, n (%) |  |  |  |  |
| White | 33 (42) | 4 (31) | 27 (55) | p = 0.101 |
| Black | 7 (9) | 3 (23) | 7 (14) |  |
| Asian | 15 (19) | 4 (31) | 7 (14) |  |
| Mixed | 2 (3) | 1 (8) | 0 (0) |  |
| Other | 21 (27) | 1 (8) | 8 (16) |  |
| **Charlson co-morbidity**<br>score, n (%) |  |  |  |  |
| 0 | 57 (63) | 4 (24) | 27 (49) | p = 0.095 |
| 1 | 25 (28) | 11 (65) | 17 (31) |  |
| 2 | 6 (7) | 1 (6) | 7 (13) |  |
| 3+ | 2 (2) | 1 (6) | 4 (7) |  |
| **ICU admission** , n (%) |  |  |  |  |
| Yes | 52 (58) | 17 (100) | 39 (72) | p = 0.015 |
| No | 38 (42) | 0 (0) | 15 (28) |  |
| **Corticosteroid use**, n (%) |  |  |  |  |
| Yes | 79 (89) | 15 (88) | 35 (65) | p = 0.076 |
| No | 10 (11) | 2 (12) | 19 (35) |  |
| **BSI organism**, n (%) |  |  |  |  |
| Gram-negative bacilli | N/A | 10 (40) | 32 (62) | p = 0.507 |
| *Enterococcus sp.* |  | 2 (8) | 12 (23) |  |
| *Staphylococcus aureus* |  | 4 (16) | 7 (13) |  |
| Other |  | 1 (4) | 1 (2) |  |

BSI = blood stream infection. \* relates to statistical comparison between COVID-19 patients who did or did not receive tocilizumab prior to the onset of BSI. Mann–Whitney test was used to compare age, Fisher’s exact test was used to compare gender and microbiology results, and Chi-square test was used to compare ethnicity and Charlson co-morbidities.

**Fig S1.**
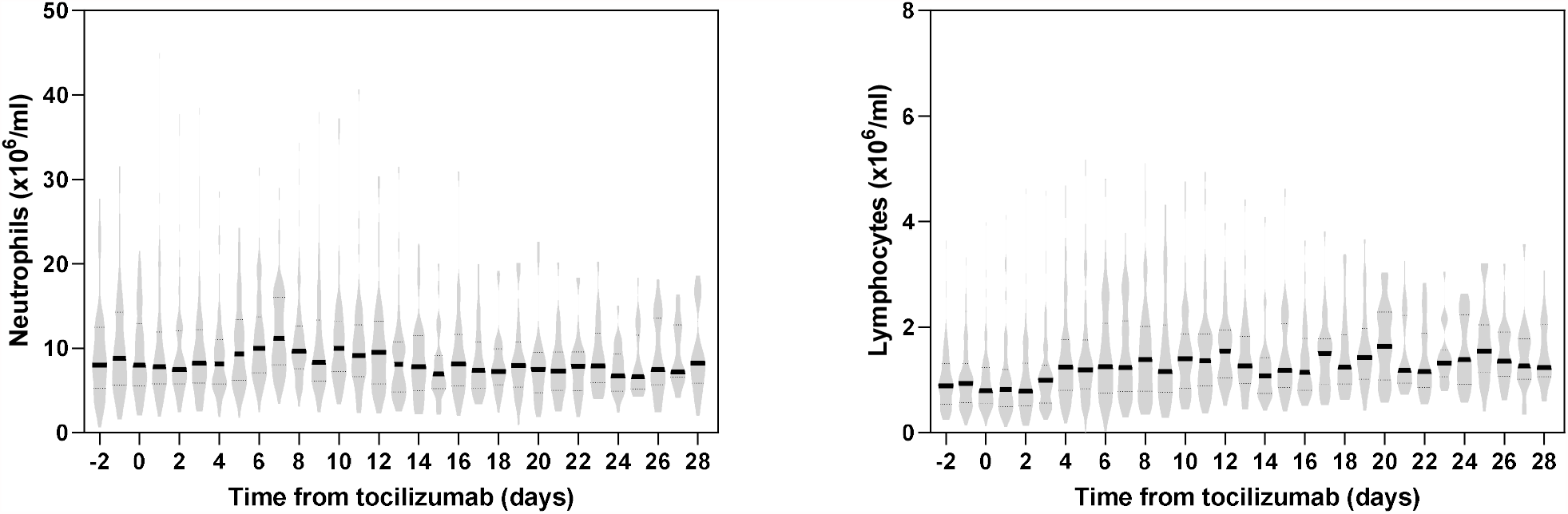
Neutrophil (left panel) and lymphocyte (right panel) counts relative to time since tocilizumab administration. All COVID-19 patients receiving tocilizumab included in analysis, independent of a subsequent presence of BSI (n = 107). Violin plots represent frequency distribution of all samples, with bold and dashed lines representing median and quartile values.

**Fig S2.**
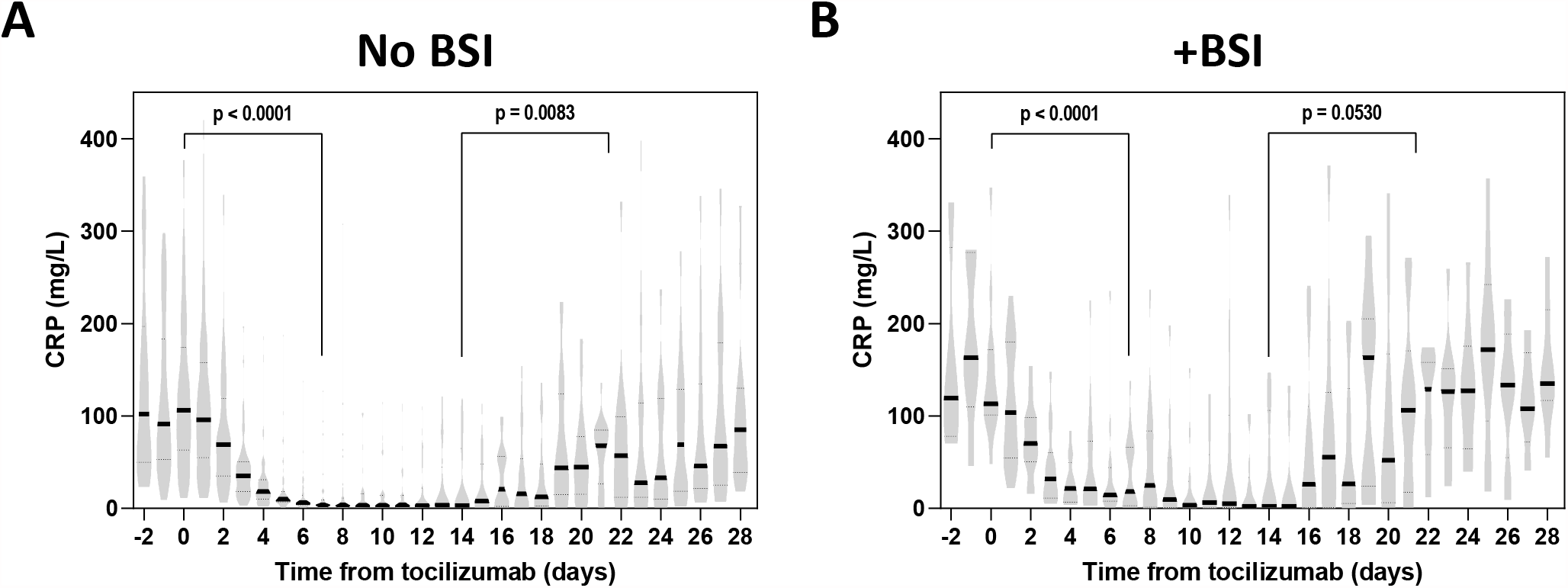
CRP concentration relative to time since tocilizumab administration in patients that (A) did not develop (n = 90) or (B) did develop a BSI (n = 17) during the same admission. Violin plots represent frequency distribution of all samples, with bold and dashed lines representing median and quartile values.

